# Evidence for secondary thrombotic microangiopathy in COVID-19

**DOI:** 10.1101/2020.10.20.20215608

**Authors:** Joseph M. Sweeney, Mohammad Barouqa, Gregory J. Krause, Jesus D. Gonzalez-Lugo, Shafia Rahman, Morayma Reyes Gil

## Abstract

The causes of coagulopathy associated with COVID-19 disease are poorly understood. We aimed to investigate the relationship between markers of endothelial activation, intravascular hemolysis, coagulation, and organ damage in COVID-19 patients and study their association with disease severity and mortality. We conducted a retrospective study of 181 hospitalized COVID-19 patients randomly selected with equal distribution of survivors and non-survivors. Patients who died had significantly lower ADAMTS13 activity, significantly higher LDH, schistocytes and von Willebrand Factor levels compared to patients discharged alive. Only 30% of patients with an initial ADAMTS13 activity <43% survived vs. 60% with ADAMTS13 ≥43% who survived. In conclusion, COVID-19 may manifest as a TMA-like illness in a subset of hospitalized patients. Presence of schistocytes on admission may warrant a work-up for TMA. These findings indicate the need for future investigation to study the relationship between endothelial and coagulation activation and the efficacy of TMA treatments in COVID-19.

## Introduction

Coronavirus disease 2019 (COVID-19) is a respiratory disease with heterogeneous manifestations ranging from asymptomatic illness in some, to systemic inflammation, multi-organ failure and a rapid death in others.^1,2^ The first stage of disease manifests as an upper respiratory infection followed by pneumonia when the virus invades the respiratory epithelium via binding to angiotensin converting enzyme 2 (ACE2) receptors.^3^ A second, more severe, phase may be manifested as multiorgan damage, including respiratory, cardiac, hepatic and renal injury. At this stage, the ACE2 receptors on the endothelium can also be involved, causing direct damage to blood vessels and inducing a coagulopathy.^3^

Systemic inflammation and coagulopathy are characteristic hallmarks of this phase. “COVID coagulopathy” manifests mainly as a prothrombotic state affecting both large and small blood vessels, and presenting as arterial, venous and microangiopathic thrombotic events.^4,5^ Coagulopathy with D-dimer elevations are reported in most hospitalized COVID-19 patients.^6-8^ A recent study showed that markers of endothelial damage such as VWF and soluble thrombomodulin were also increased in COVID-19 hospitalized patients. All these markers were even higher in intensive care unit patients and correlated with mortality.^9^ VWF has three main functions: binding to collagen in the wounded subendothelial matrix, binding to glycoprotein-1b on platelets and carrying/delivering coagulation factor VIII (FVIII) to the surface of activated platelets bound to wounded endothelium.^10^ Whether the increased VWF reported in COVID-19 is a result of increased production, abnormal and/or increased release, or decreased destruction is unclear. Since ADAMTS13 (a disintegrin and metalloproteinase with a thrombospondin type 1 motif, member 13), a von Willebrand factor-cleaving protease, plays a key role in regulating both VWF quantity and multimer size, analysis of this enzyme would be important in elucidating the pathophysiology of COVID coagulopathy. Although there have been some COVID-19 data involving ADAMTS13 levels, the small sample size in these reports precluded any major conclusions.^11-13^

The primary objective of our study was to establish the relationship of endothelial activation and VWF-related biomarkers with coagulation/thrombosis, intravascular hemolysis and end-organ damage in a large cohort of COVID hospitalized patients. The secondary objective was to study the correlation of VWF-related biomarkers with disease severity and mortality.

## Methods

### Study Population

We included confirmed COVID-19 cases in Montefiore Medical Center who were hospitalized and had routine and/or advanced coagulation tests done between March 26th to May 5th, 2020. All included patients tested positive for SARS-CoV2 with reverse-transcriptase–polymerase-chain-reaction real-time assay of the nasal and the pharyngeal swabs. We excluded the patients younger than 18 years of age. The medical records of the patients were reviewed to obtain epidemiological, demographic, clinical and laboratory data. The management and clinical outcomes were followed-up until June 20, 2020. All cases had final disposition (expired or discharged alive) and none were censored. The study was approved by the Albert Einstein College of Medicine Institutional Review Board.

### Laboratory Investigations

Plasma samples were aliquoted from sodium citrate tubes shortly after collection and stored −80°C until needed. To study the association of disease severity with VWF and ADAMTS13, we randomly selected 181 plasmas with equal numbers of discharged and deceased patients across the range of D-Dimer levels from normal to very high (0.26 to >20 μg/mL). To study the association between platelet count and ADAMTS13 activity, we selected 13 of those 181 samples from patients who had a platelet count <70 x10^6^/uL on admission. The cut off <70 x10^6^/uL was predefined, as it is a common cut off to differentiate mild from moderate thrombocytopenia in our institute.

### Statistical Analysis

Data analysis was performed using R studio, V.3.6.2 and graphs generated in Prism V.8.3.1. Differences in demographic, clinical variables and laboratory assessments between patients who were deceased and patients discharged alive were compared using chi-square or Fisher’s exact tests for categorical variables, two-sample Student t tests, and one-way ANOVA for three group comparisons. Youden’s *J* statistics was used to find the optimal cutpoint for ADAMTS13 for mortality. Logistic regression of initial ADAMTS13 adjusted by age was represented in a density plot against mortality. A Kaplan Meier cumulative curve was generated for patients discharged alive.

### Laboratory Testing

Coagulation tests (VWF Antigen, VWF Ristocetin activity, FVIII levels, D-Dimer, and fibrin monomer (FM) were performed by STA-R Max instruments using STAGO reagents as per manufacturer recommendations. STA Liatest LIA D-Dimer assay was performed as per manufacturer recommendations and reported as FEU μg/mL. Complete blood counts were performed by Sysmex XN9000. Chemistry assays were performed by Roche instrumentation and reagents as per manufacturer recommendations.

After thawing the patient samples in a 37 °C water bath, a 1:1 dilution was created using patient plasma and STAGO Owren-Koller buffer. Using this dilution, VWF Antigen, VWF ristocetin cofactor activity, FVIII activity levels, D-Dimer, and FM levels were obtained. If the STA-R Max instrument was not able to accurately report the value due to it being above the reportable range, a 1:20 dilution was made with Owren-Koller buffer and subsequently a 1:100 dilution until the value was within reportable range.

### Data Gathering

Chart reviews were performed to document demographic attributes (age, sex and self-reported race and/or ethnicity) and baseline comorbidities (body mass index, previous history of hypertension, diabetes, kidney, pulmonary, liver, autoimmune, cancer, or sickle cell disease) on presentation collected for calculation of comorbidity Indexes. We gathered data on initial vital signs and laboratory values within the first 48 hours of hospital admission. The laboratory assessments consisted of a complete blood count, blood chemical analysis, coagulation testing, assessment of liver and renal function, measures of electrolytes, and markers of inflammation. Additionally, we noted the D-Dimer, fibrinogen, hemoglobin, creatine, LDH, indirect bilirubin. and platelet count of the patients within 48 hours of the time of collection of the samples we tested. We also noted the trajectory of these parameters in the week following the collection of the sample, noting whether these parameters increased by 10% or more, decreased by 10% or more, or remained stable. We accessed each patient for thrombosis and clot formation. A patient was considered to have thrombosis if a thrombus was identified on radiological imaging. We also noted if a patient experienced *ex vivo* clotting while on hemodialysis (HD) or continuous renal replacement therapy (CRRT). We documented anticoagulation medications given to each patient within 48 hours preceding the thrombus or clotting event.

### STROBE Criteria

This study followed the STROBE criteria for retrospective studies including: 1) providing a summary in the abstract of the objectives, the study type, outcome and conclusion; 2) providing scientific background, rationale and hypothesis in the introduction; 3) providing details of the study design in the methods including setting, participants, sample size, variables, data sources and measurements; 4) providing details of statistical methods; 5) describing demographics of the population in the results; 6) providing 95% confidence intervals in the results when appropriate; 7) providing a discussion of the limitations, potential bias and generalizability.

### VWF Multimers

VWF multimers were generated using a western blot technique previously described. ^14^ Briefly, samples were first prepared by normalizing the amount of thawed plasma added to the loading dye according to the measured VWF antigen level. The samples were then heat inactivated for at least 15 minutes in a 56 °C water bath. The samples subsequently were loaded onto a 1% agarose gel. A normal control consisting of normal pooled plasma and an abnormal control consisting of plasma from a patient with Type 2A Von Willebrand Disease were used. The gels were ran using the PhastSystem Separation and Control unit. After this, the gels were transferred to a polyvinylidene difluoride membrane also using the PhastSystem Separation and Control unit. After the transfer, the membranes were blocked for at least 40 minutes using 1.2% Bovine Serum Albumin (BSA) (Sigma-Aldrich) in tris-buffered saline plus 0.04% Tween 20. After blocking, rabbit anti-human F. VIII related antigen antibody (Accurate Chemical AXL 205) was applied as the primary antibody for 45 minutes at a concentration of 5.58 µg/mL diluted in 1.2% BSA. Anti-rabbit IgG-alkaline phosphatase-conjugated antibody (Sigma A8025) was applied as the secondary antibody for 45 minutes at a concentration of 1.5 µg/mL diluted in 1.2% BSA. After this, SigmaFast 5-Bromo-4-chloro-3-indolyl phosphate/nitro blue tetrazolium (BCIP/NBT) solution was used to develop the blot.

Western blots were then analyzed using the ImageJ analysis software. Briefly, each sample well was segmented into low, intermediate, and high molecular weight VWF multimers. Low molecular weight multimers were defined as the bottom three bands of the well. The division between intermediate and high molecular weight multimers was established using the abnormal control that was run in each gel, as the abnormal control has no high molecular weight multimers. A straight line was drawn across the gel where the high molecular weight multimer signal in the abnormal control started to taper. Intermediate size multimers were those between the cut-off established with the abnormal control and the highest band of the low molecular weight multimers. The measure function in ImageJ was used to measure the raw integrated density of each size of the multimers. All values were normalized to total VWF protein loaded per well. Values are reported as fold change from normal control.

### VWF Collagen Binding Activity

Previously thawed plasma was centrifuged for 10 minutes at 24,328xg and the supernatant was used for the VWF collagen binding activity ELISA and ADAMTS13 Activity Assay. Human von Willebrand Factor: collagen binding activity was measured using Zymutest vWF:CBA ELISA Kit (#RK038 from Hyphen BioMed). The ELISA was completed and analyzed using the manufacturer’s recommendations.

### ADAMTS13 Activity

The ADAMTS13 protease activity on previously thawed and centrifuged plasma supernatant was measured using ATS-13 Activity Assay based on fluorescence resonance energy transfer (Immucor ATS-13). The assay was carried out and analyzed using the manufacturer’s recommendations. For patients whose plasma samples had an ADAMTS13 activity level of less than 30%, we ran an inhibitor assay according to the manufacturer’s recommendations. Briefly, this assay involved mixing equal volumes of normal pooled plasma with the patient’s plasma and measuring the ADAMTS13 activity of the mixed sample relative to that of the normal pooled plasma to find the percent inhibition. We considered a value greater than 40% inhibition to indicate a true inhibitor.

### ADAMTS13 Antigen

The ADAMTS13 antigen level was measured on previously thawed and centrifuged plasma supernatant using Technozym® ADAMTS-13 Fluorogenic Activity/Antigen (cat# 5450551). The ELISA was completed and analyzed using the manufacturer’s recommendations.

### ADAMTS13 Antibody Detection

The presence of human IgG autoantibodies against ADAMTS13 was determined using Technozym® ADAMTS-13 Inhibitor ELISA (cat# 5450451). The ELISA was completed and analyzed using the manufacturer’s recommendations.

### Peripheral blood smear

CBCs were performed on admission and when clinically indicated during hospitalization. Manual differentials were performed when reflexed due to a count threshold or scattergram abnormality. We analyzed all the smears available in which a smear review was reflexed due to an abnormality in the WBC, RBC, platelet count or scattergram. Smear photos were obtained from CellaVision. Schistocytes and ghost cell count was based on at least 1000 RBCs. RBC count was performed by using the digital manual counter on Image J. We noticed that cases in which the schistocytes were less than 1%, the smear review was not prompted by RBC or platelet flags but were prompted by unrelated flags, e.g. WBC flags. As previously reported, the sensitivity of RBC or platelet flags to detect schistocytes is less than 1% (0.6-0.9%). ^15^ Thus, we classified all cases with no RBC/platelet flags as <1% schistocytes.

## Results

### Study Population

Samples from 90 patients who died and 91 who were discharged alive with a wide and balanced distribution of D-Dimer levels (0.26 to >20 μg/mL) were selected. The characteristics of the study population are summarized in Table 1. Consistent with many other studies, non-survivors were older (median [interquartile range (IQR)]; 72.5 [63.3, 79.8] vs 62.0 [50.5, 70.0] years) and the majority were male (67% (60/90), p=0.03) (Table 1). No difference between survivors and non-survivors by ethnicity or comorbidity was observed. As expected, the number of patients that required a ventilator was higher in non-survivors 51% (46/90) vs. 26% (24/91) in survivors. No significant difference in the average length of stay or treatment was observed.

**Table 1).** Characteristics and Initial Clinical Laboratory Data of 181 Patients with COVID19 Stratified by Mortality Outcome

| Characteristics |  | Discharged (n=91<sup>a</sup>)<br>(median [IQR] or n (%)) | Deceased (n=90<sup>a</sup>)<br>(median [IQR] or n (%)) | p |
| --- | --- | --- | --- | --- |
| Age |  | 62.0 [50.5, 70.0] | 72.5 [63.3, 79.8] | <0.001 |
| Sex (male) |  | 46 (51) | 60 (67) | 0.03 |
| Race | Black or African-American | 36 (40) | 34 (38) | 0.81 |
|  | White | 9 (10) | 11 (12) | 0.81 |
|  | Other/Patient Declined/ Not reported | 46 (51) | 44 (49) | 0.82 |
| Ethnicity | Spanish/Hispanic/Latino | 36 (40) | 33 (37) | 0.69 |
| BMI (kg/m<sup>2</sup>) |  | 30.1 [27.2, 34.0] | 28.5 [25.5, 31.8] | 0.04 |
| Elixhauser Comorbidity Index |  | 4.0 [1.0, 6.3] | 5.0 [2.0, 8.0] | 0.14 |
| Length of Stay (days) |  | 9.0 [5.0, 27.0] | 10.5 [5.3, 16.0] | 0.73 |
| In Vivo Thrombosis or Ex Vivo Clot |  | 23 (25) | 20 (22) | 0.63 |
| Invasive Ventilator Use |  | 24 (26) | 46 (51) | <0.001 |
| Vasopressor Use |  | 21 (23) | 31 (34) | 0.09 |
| Hemodialysis or CRRT Use |  | 17 (19) | 23 (26) | 0.27 |
| Anticoagulation Use<sup>b</sup> |  | 34 (37) | 33 (37) | 0.92 |
| Steroid Use<sup>c</sup> |  | 29 (32) | 31 (34) | 0.71 |
| Initial Clinical Laboratory Values Measured upon Admission (units) [Reference Range] |  |  |  |  |
| eGFR (mL/min/1.73m<sup>2</sup>) [90-120] |  | 74.5 [34.3, 98.8] | 40.5 [25.6, 70.8] | <0.001 |
| Creatinine (mg/dL) [0.84-1.21] |  | 1.0 [0.7, 1.9] | 1.7 [1.1, 2.5] | <0.001 |
| Anion Gap (mEq/L) [8-16] |  | 17.0 [15.0, 20.0] | 18.0 [16.0, 22.0] | 0.03 |
| Aspartate Transaminase(IU/L) [8-48] |  | 44.0 [28.3, 68.0] | 51.0 [37.0, 78.8] | 0.08 |
| Neutrophil to Lymphocyte Ratio [0.78-3.53] |  | 5.9 [3.0, 9.0] | 7.1 [4.6, 11.3] | 0.02 |
| Platelet (Count) (k/uL) [150-450] |  | 213.0 [157.3, 288.0] | 183.0 [136.0, 270.0] | 0.14 |
| D-Dimer (ug/mL) [<.27] |  | 1.8 [0.7, 3.9] | 2.6 [1.3, 5.7] | 0.03 |
| International Normalized Ratio [<1.1] |  | 1.1 [1.0, 1.2] | 1.1 [1.0, 1.3] | 0.65 |
| C-Reactive Protein (mg/L) [<10] |  | 12.4 [4.9, 20.3] | 15.9 [6.9, 26.1] | 0.06 |
| Lactate Dehydrogenase (U/L) [140-280] |  | 451.0 [277.0, 658.0] | 542.0 [391.0, 652.0] | 0.03 |
| Troponin (ng/mL) [<0.04] |  | 0.01 [0.01, 0.02] | 0.03 [0.01, 0.06] | <0.001 |
| Pulse Oximeter (%) [95-100] |  | 96.0 [91.0, 99.0] | 94.0 [87.3, 97.8] | 0.04 |
| Diastolic Blood Pressure (mm Hg) [60-80] |  | 75.0 [66.0, 85.0] | 62.0 [42.8, 75.0] | <0.001 |

a. Unless otherwise stated
b. within 48 hours before clot or ADAMTS13 measurement
c. within 24 hours before ADAMTS13 measurement
Abbreviations: IQR, interquartile range; BMI, body mass index; CRRT, Continuous Renal Replacement Therapies; eGFR, Estimated Glomerular Filtration Rate

### Initial Clinical Laboratory Data

Initial markers of renal function were significantly worse in non-survivors compared to survivors (creatinine, 1.7 [1.1, 2.5] vs 1.0 [0.7, 1.9] mg/dL, p<0.001) whereas markers of liver function were not significantly different. Oxygen saturation was lower in non-survivors compared to survivors (94.0 [87.3, 97.8] vs 96.0 [91.0, 99.0] %; p=0.04). Also, initial diastolic blood pressure was significantly lower in non-survivors vs. survivors (62.0 [42.8, 75.0] vs 75.0 [66.0, 85.0] mmHg; p<0.001). CBC parameters were not significantly different with the exception of neutrophil to lymphocyte ratio, (7.1 [4.6, 11.3] in non-survivors vs 5.9 [3.0, 9.0] in survivors; p=0.02).

The only hemolysis marker that was significantly higher in non-survivors was lactate dehydrogenase (LDH) (542.0 [391.0, 652.0] vs. 451.0 [277.0, 658.0] U/L; p<0.001). Initial D-Dimer was significantly higher in non-survivors (2.6 [1.3, 5.7] vs. 1.8 [0.7, 3.9] μg/mL; p=0.03) (Table 1).

### Endothelial and coagulation activation

We analyzed ADAMTS13 activity, VWF, FVIII, D-Dimer and FM, a precursor of D-Dimer, on the same samples irrespective of the time since admission. Non survivors had significantly lower ADAMTS13 activity levels (48.8 [36.2, 65.1] vs. 63.6 [47.2, 78.9]%; p=<0.001) and higher FM (13.2 [5.0, 129.1] vs. 5.0 [5.0, 29.40]μg/mL; p=0.02) and D-Dimer levels (4.93 [1.83, 20.00] vs 2.90 [0.92, 14.47]ug/mL; p=0.04) than survivors (Table 2). As expected, VWF antigen directly correlates with VWF activity Ristocetin Cofactor (VWF:RCo) (r=0.58; p=<0.0001) and VWF:RCo correlates with VWF activity collagen binding (VWF:CB) (r=0.77; p=<0.0001) and VWF antigen correlates with FVIII (r=0.34; p=<0.001) (Supplementary Figure1A-C). Also as expected, ADAMTS13 activity inversely correlates with VWF:RCo (r=-0.28; p=0.0001) and VWF:CB (r=-0.3; p=0.009) (Supplementary Figure1D-E).

**Table 2).** Markers of endothelial activation, intravascular hemolysis, coagulation, and organ damage of 181 Patients with COVID19 Stratified by Mortality Outcome

| Clinical Laboratory Values (units) [Reference Range] | Discharged (n=91<sup>a</sup>)<br>(median [IQR] or n (%)) | Deceased (n=90<sup>a</sup>)<br>(median [IQR] or n (%)) | p |
| --- | --- | --- | --- |
| ADAMTS13 Activity (%) [70-110] | 63.6 [47.2, 78.9] | 48.8 [36.2, 65.1] | <0.001 |
| Schistocyte Count (%) [<0.5] | 0.56 [0.16, 1.12] n=31 | 1.06 [0.49, 2.63] n=42 | 0.008 |
| Schistocyte >1% | 10 (11) | 22 (24) | 0.02 |
| Lactate Dehydrogenase (U/L) [140-280] | 424.0 [275.0, 594.0] | 562.5 [437.3, 664.8] | <0.001 |
| Indirect Bilirubin (mg/dL) [0.2-0.8] | 0.2 [0.1, 0.4] | 0.2 [0.1, 0.4] | 0.89 |
| Hemoglobin (g/dL) [12-17.5] | 11.8 [10.1, 13.9] | 11.7 [9.7, 13.7] | 0.85 |
| Platelet Count (k/uL) [150-450] | 241.0 [163.5, 344.5] | 196.0 [124.8, 312.8] | 0.06 |
| Decreased Platelet Trajectory<sup>b</sup> | 18 (20) | 27 (30) | 0.11 |
| Creatinine (mg/dL) [0.84-1.21] | 1.0 [0.7, 2.3] | 1.9 [1.2, 3.5] | <0.001 |
| Increased Creatinine Trajectory<sup>b</sup> | 14 (15) | 37 (41) | 0.001 |
| VWF Activity (Ristocetin) (%) [50-150] | 282.0 [214.0, 400.0] | 321.0 [238.0, 451.0] | 0.05 |
| VWF Activity (Collagen Binding) (%) [50-150] | 383.2 [235.2, 458.2] n=35 | 368.7 [261.1, 585.2] n=37 | 0.40 |
| VWF Antigen (%) [50-150] | 362.0 [261.0, 540.0] | 441.0 [307.6, 598.0] | 0.05 |
| D-Dimer (ug/mL) [<.27] | 2.90 [0.92, 14.47] | 4.93 [1.83, 20.00] | 0.04 |
| Fibrin Monomer (ug/mL) [<10] | 5.00 [5.0, 29.40] | 13.2 [5.0, 129.1] | 0.02 |
| Factor VIII Activity (%) [50-150] | 175.0 [118.0, 247.5] n=27 | 160.0 [106.5, 246.5] n=58 | 0.46 |
| HMW Multimer Fold Change from Normal<sup>c</sup> | 0.95 [0.71, 1.25] n=57 | 1.04 [0.83, 1.46] n=58 | 0.14 |
| Fibrinogen (mg/dL) [200-400] | 572.0 [462.0, 727.0] n=59 | 496.0 [376.3, 744.3] n=50 | 0.27 |

a. Unless otherwise stated
b. Based on a change >10% over 1 week period
c. See figure 2
Abbreviations: IQR, interquartile range; ADAMTS13, a disintegrin and metalloproteinase with a thrombospondin type 1 motif, member 13; VWF, Von Willebrand Factor; HMW, High Molecular Weight

Thus, we analyzed the trends of ADAMTS13 and VWF levels stratified by D-Dimer. When stratified by D-Dimer <2, 2-10, >10 μg/ml, ADAMTS13 levels incrementally decrease with higher D-Dimer (Supplementary Figure 1G). These D-Dimer cut offs were based on our previous studies of D-Dimer correlation with mortality (Billett et al., accepted Thrombosis and Haemostasis). Likewise, VWF:RCo and VWF antigen incrementally increase based on D-Dimer levels (Supplementary Figure 1H-I). In addition, similar trends are seen when VWF:CB and VWF:RCo were stratified by FM (Supplementary Figure 1K-L).

### Increased high molecular weight multimers in COVID-19 inpatients

VWF multimer analysis was performed in the first 115 samples analyzed. We observed that many COVID-19 patients had an increased density of high molecular weight multimers (HMWM) compared to normal pooled plasma (Table 2, Figure 1a). Increased HMWM correlated with higher VWF:RCo (r=0.5; p<0.0001, Supplementary Figure 1F) and increasing D-Dimer (p<0.01, Supplementary Figure 1J). However, the relative increased HMWM was not significantly different between survivors and non-survivors (Table 2). Therefore, no further VWF multimer analysis was performed in the remaining cases. Nonetheless, serial time points in a patient showed how HMWM changed along with VWF levels and inversely with ADAMTS13 levels (Figure 1B, D).

**Figure 1).**
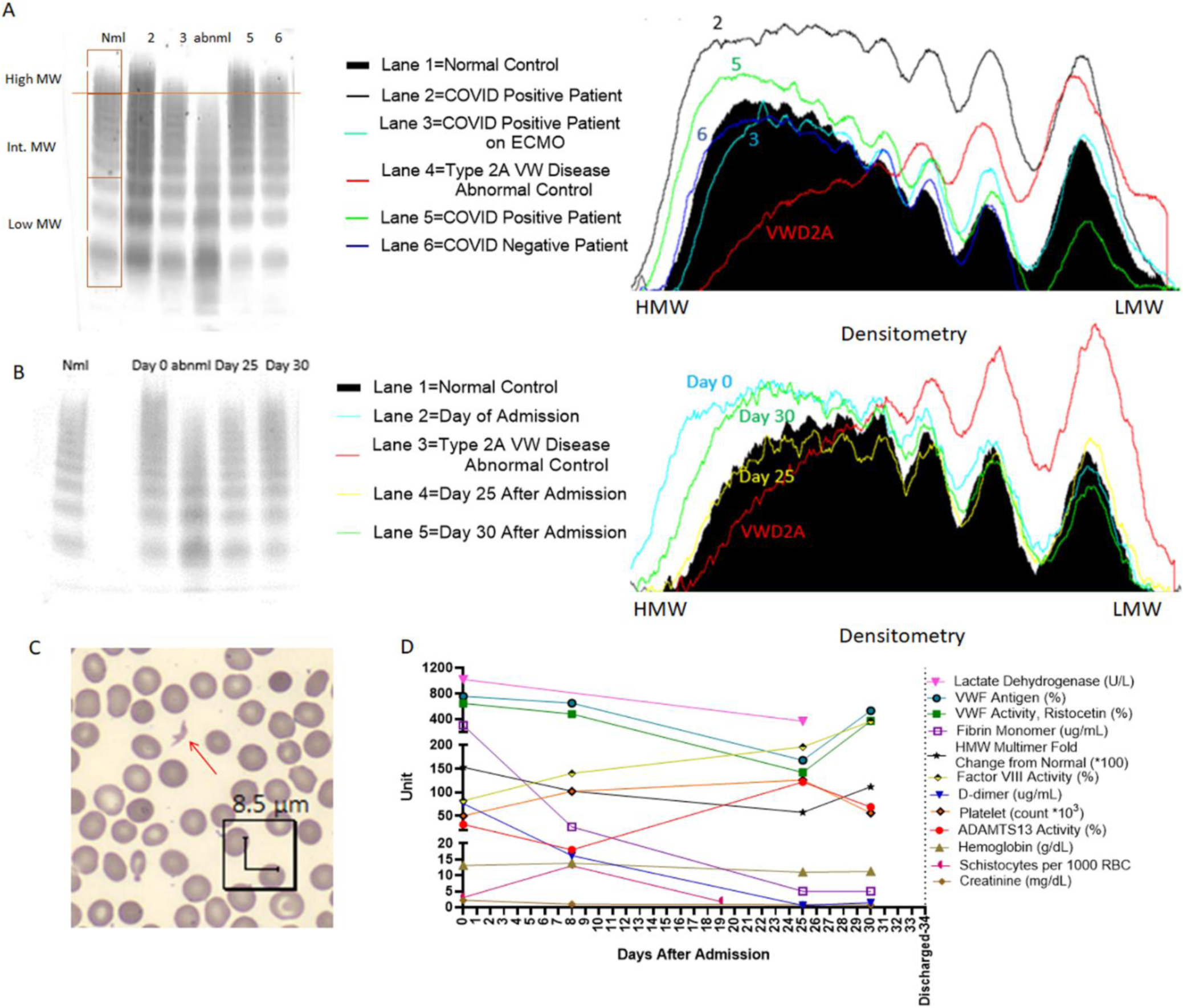
Cross sectional and longitudinal analysis of VWF Multimers and other markers of endothelial activation, hemolysis, and coagulation. For each multimer western blot, patient plasma was run on each lane, and the loading of all samples was normalized to measured VWF antigen levels. For each western blot, Bands 1-3 were considered low Molecular Weight Multimers, the bands between band 4 and the last band of the abnormal control were considered intermediate molecular weight multimers, and the bands above the last band of the abnormal control were considered high molecular weight multimers A) The Western Blot to the left shows a pattern of VWF multimer cleavage in five patients. Lane 1 is the negative control, which was derived from normal pooled plasma, and lane 4 is the abnormal control, which was derived from the plasma of a patient with Type II von Willebrand disease. The abnormal control is missing high molecular weight multimers. Lanes 2, 3, and 5 are from COVID-19 positive patients. The patient in lane 6 is from a COVID-19 negative patient with a normal multimer pattern. The COVID-19 positive patients have increased high molecular weight multimers, except for the patient in lane 3 who was on extracorporeal membrane oxygenation at the time of sample collection. The image to the right of the blot is the densitometry of the lanes represented in the western blot. The black filled in area represents the density of the normal control, and the red line indicates the abnormal control. The other lines indicate the densitometry of the multimers of the patients. B) Longitudinal Trends of Coagulation Parameters of a Discharged COVID-19 Patient. Western blot on the left shows the change in HMW multimer patterns throughout this patient’s hospital stay. Lane 1 is the negative control and Lane 3 is the abnormal control. Multimer pattern between lane 1 and 2 was blocked as it was derived from an unrelated COVID-19 patient. Lane 2 corresponds to the day of admission, Lane 4 corresponds to day 25 after admission, and Lane 5 corresponds to day 30 after admission. The image to the right of the blot is the densitometry of the lanes represented in the western blot. The black filled in area represents the density of the normal control, and the red line indicates the abnormal control. The other lines indicate the densitometry of the multimers at various timepoints of the patient. Note that by day 25, HMW multimers decreased to normal size. C) An example of a schistocyte (red arrow) from a blood smear from day 0 from the patient in panel B. D) Line graph showing various coagulation, endothelial activation and hemolysis parameters throughout the hospital course of the patient from panel B until the patient’s discharge on day 34 after admission. Many parameters were abnormal upon admission. Particularly, on day 8, when the patient had the lowest recorded ADAMTS13 activity (18%), many parameters were abnormal, including an elevated schistocyte count and VWF levels. All parameters tended to improve throughout the patient’s hospital stay and became closer to normal by day 25.

### Increased schistocytes and LDH are associated with low ADAMTS13 and higher mortality

Upon smear review, many RBC and platelet abnormalities were observed including fibrin strands, platelet clumps, giant platelets, echinocytes, elliptocytes, ghost cells, tear drops, schistocytes, and RBC agglutination (Figure 2). Importantly, schistocytes along with microspherocytes were among the most remarkable and predominant findings (Figure 1C, Figure 2). Increased percentage of schistocytes correlated with high VWF levels (r=0.24; p=0.04) (Supplemental Figure2A-C). Increased percentage of schistocytes also correlated with decreased platelet count (r=-0.26; p=0.02) and low ADAMTS13 activity (r=-0.45; p<0.0001) (Supplemental Figure 2D-F). Increased percentage of schistocytes correlated with markers of hemolysis, such as LDH (r=0.51; p<0.0001) and indirect bilirubin (Supplemental Figure 2G-I). Similarly, increased LDH strongly correlated with high VWF:RCo (r=0.25; p=0.002) and VWF antigen (r=0.34; p<0.0001) levels (Supplemental Figure 3A-B). Importantly, LDH strongly correlated with indirect bilirubin (r=0.46; p<0.0001), supporting their use as hemolysis markers (Supplemental Figure 3C). High LDH correlated with increasing creatinine (r=0.16; p<0.05), and creatinine also correlated inversely with hemoglobin (r=-0.18; p=0.02) and trended with decreasing platelet count (r=-0.14; p=0.06) (Supplemental Figure 3D-F). The percentage of schistocytes was higher in those who died than those who survived (1.06 [0.49, 2.63] vs. 0.56 [0.16, 1.12], p=0.008) (Table 2).

**Figure 2).**
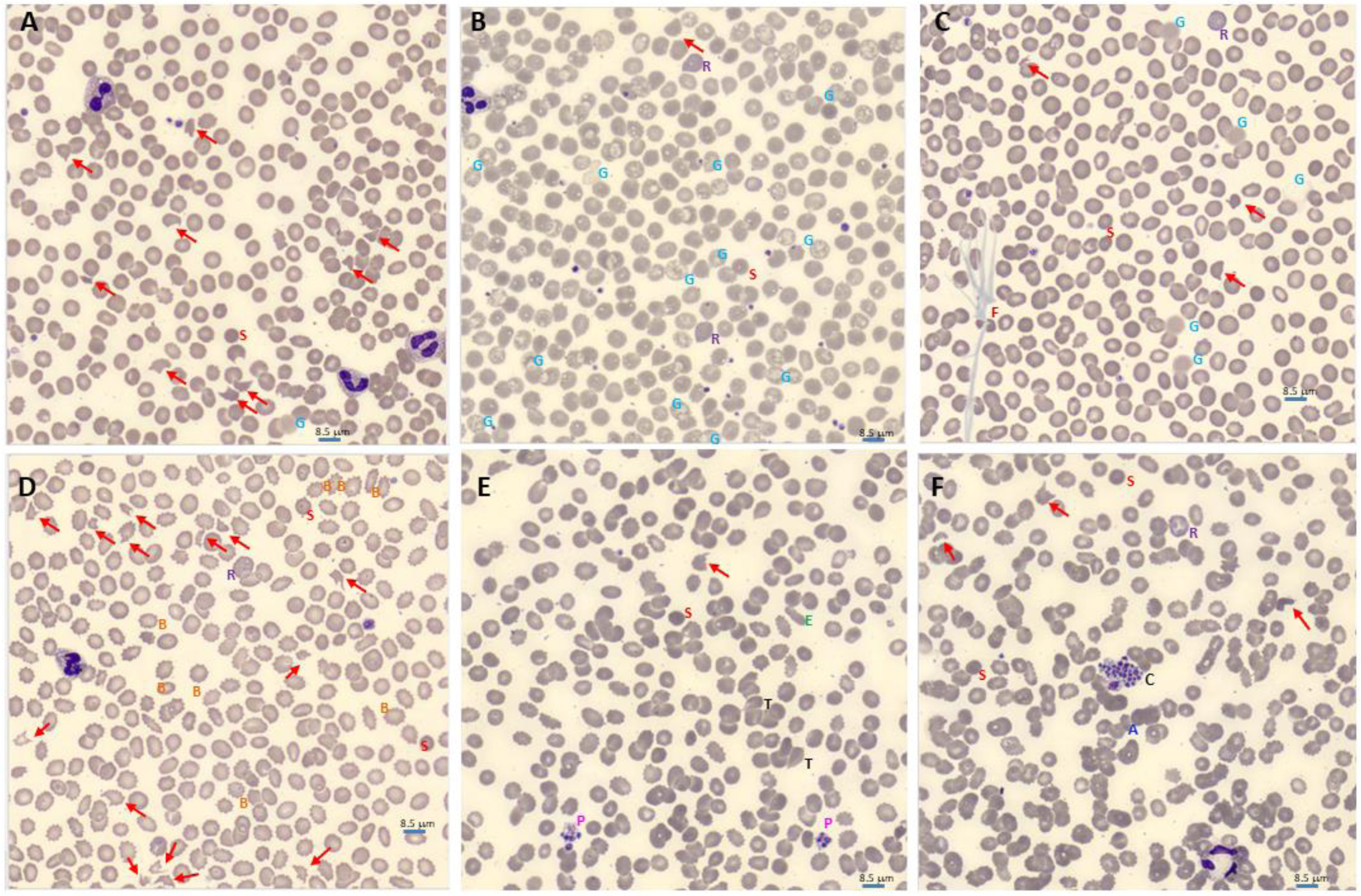
Blood smear abnormalities in COVID-19. Morphology evaluation of peripheral blood smear were performed by reviewing digital images from CellaVision®. Schistocytes (red arrow) was a predominant finding in many COVID-19 patients. Spherocytes and microspherocytes (red S) were very abundant, especially in smears with high number of schistocytes, thus only one is pointed in each smear for illustration (A-F). Ghost red blood cells (blue G) was also a common finding. Several morphologies of ghost cells were seen: smudge and diffuse hemoglobin staining without intact membrane (A and C), vacuolated red blood cells with intact membrane and vacuolated ghost without intact membrane (B). Reticulocytes (purple R) were also seen (B, C, D, F). Fibrin strands (brown F) were seen in several patients (C). Echinocytes, aka Burr cells (orange B), were seen in association with renal injury (D). Giant platelets (pink P) were seen in many COVID19 patients (E). Elliptocytes (green E) and tear drops (black T) were seen in several smears (E). Platelet clumps (Black C) and agglutination (blue A) were also seen in some in COVID19 patients (F). Scale bar = 8.5 um.

### Thrombosis, coagulation activation and VWF

We documented thrombosis, type of thrombosis, anticoagulation and temporal relationship of thrombosis to ADAMTS13 testing (Supplemental Table 1). 19% (34 of 181) patients developed *in vivo* thrombosis during hospitalization. Patients with thrombosis exhibited significantly higher D-Dimer (mean difference, 52.9; 95% CI 27.3-78.6 μg/mL), FM (517.4;168.8-865.9 μg/mL), VWF activity (62.3; 2.9-127.4%), LDH (314.5;56.4-573.0 U/L) and creatinine (1.36;0.24-2.49 mg/dL) (Supplemental Figure 4). A significant number of patients did not receive anticoagulation within 48 hours before clot detection (18 of 43 (42%)) (Supplemental Table 1). Although the number of patients with documented thrombosis (both *in vivo* and *ex vivo*) was not significantly different based on ADAMTS13 levels, *ex vivo* clots, such as clots in the hemodialysis lines, were mainly observed in patients with ADAMTS13 levels lower than the normal range <70% (10 patients (7.8%) vs. 1 (1.9%); p = 0.181). Anticoagulation did not seem to change the risk of these thrombosis (Supplemental Table 1). However, VWF antigen and LDH levels were higher among the patients that received anticoagulation (mean difference, 105.9; 95% CI 13.4-198.4%, and 239.9; 54.0-425.8 U/L, respectively) (Supplemental Figure 5).

### Initial ADAMTS13 predicts hospital course and discharge outcome

Given that ADAMTS13 levels seem to fluctuate during the course of hospitalization as shown in Figure 1D, we studied whether the initial ADAMTS13 within 72 hours of admission is predictive of mortality. 102 patients had ADAMTS13 levels performed within 72 hours of admission (Table 3). Using Youden’s *J* statistic, we determined that the best cut-off of initial ADAMTS13 to predict mortality was 43%, p-value <0.01 (Figure 3a). As expected, the demographic and clinical characteristics were similar to the larger original cohort (Table 1). There was no difference by age or gender, although Hispanics represented 49% of patients with ADAMTS13 levels <43% (Table 3). The logistic regression model of ADAMTS13 adjusted by age as a continuous variable showed that patients presenting with lower ADAMTS13 levels had higher risk of mortality (Figure 3b). Only 30% (10/33) of patients with an ADAMTS13 activity <43% within 72 hours of admission survived compared to 60% (41/69) with ADAMTS13 ≥43% who survived (Figure 3c). Patients presenting with low ADAMTS13 (<43%) had significantly higher VWF:RCo activity (352.00 [225.00, 490.00] vs. 258.00 [200.00, 322.00]%; p=0.04). The number of patients that required ventilation with an initial ADAMTS13 <43% was more than twice that of patients with initial ADAMTS13 ≥43% (12/33 (36%) vs. 11/69 (16%); p=0.04) (Table 3).

**Figure 3).**
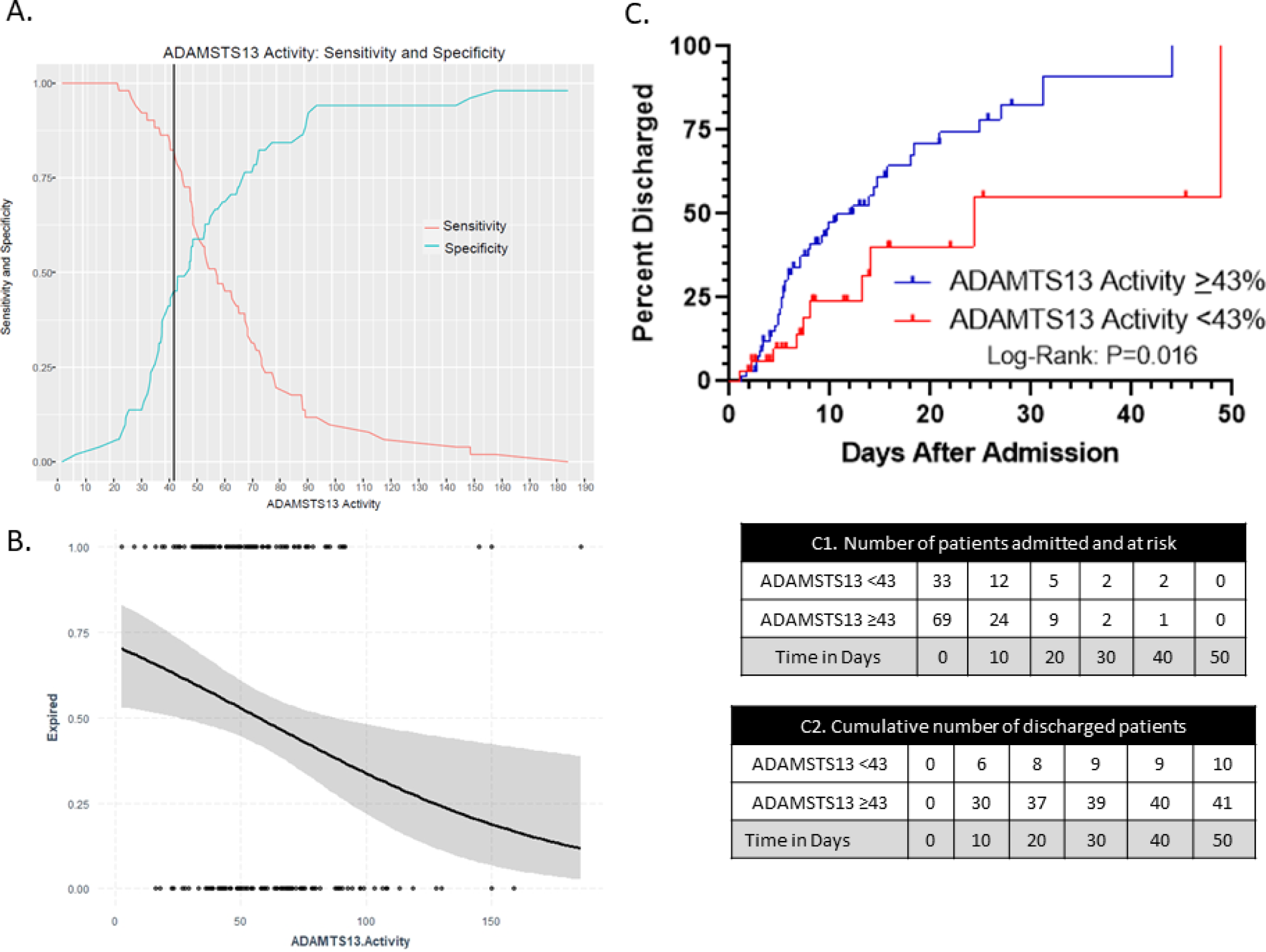
Optimal cut off of ADAMTS13 by Youden’s *J* statistics and cumulative discharged curve. A) Youden index measuring the optimal cut point for ADAMTS13 activity as a differentiating marker when equal weight is given to sensitivity and specificity for the values in the cohort. The optimal cut-point for the initial ADAMTS13 within 72 hours since admission to predict mortality was found to be at 43% with an accuracy of 0.63, sensitivity of 0.82 and AUC of 0.63. B) Logistic regresion model of initial ADAMTS13 adjusted by age. Patients that expired (each dot at the top classified as event 1) presented with lower ADAMTS13 levels compared to patients that were discharged alive (each dot at the bottom classified as event 0). The gray zone represents 95% the confidence interval. Kaplan Meier curve shows cumulative number of discharged COVID-19 positive patients over time (n=102) based on initial ADAMTS13. Patients with ADAMTS13≥43 show higher rate of discharge alive compared to patients with ADAMTS13<43 (log rank, p = 0.016). C1 table shows the number of COVID-19 positive patients admitted and at risk of mortality over time. C2 table shows the cumulative number of discharged patients in each group in increments of every 10 days. Each dot represents a discharged patient.

**Figure 4).**
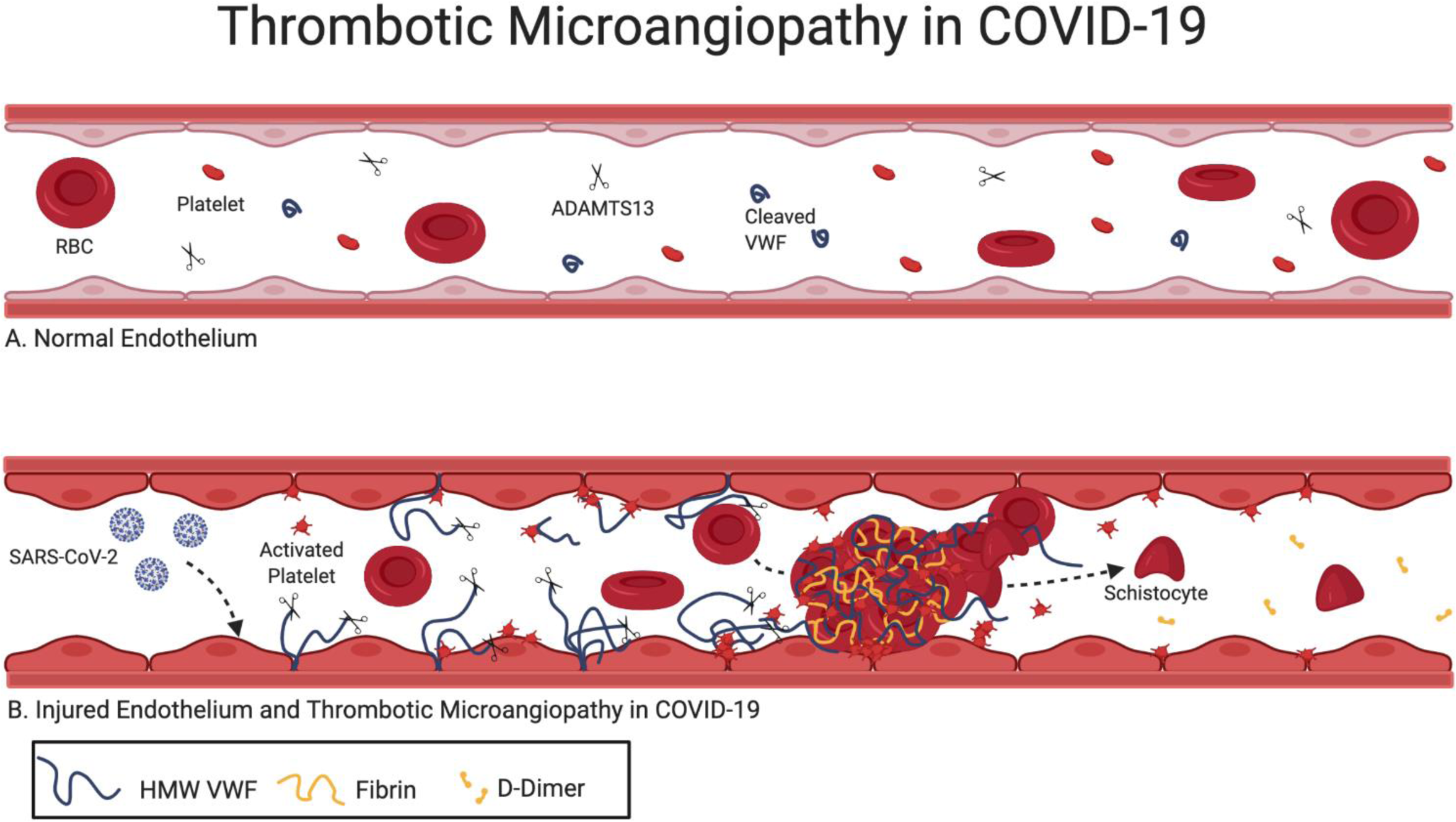
A. Normal endothelium B. SARS-CoV-2 enters the endothelial cells of capillaries via the ACE2R. Injured endothelial cells releases high molecular weight multimers VWF multimers (HMW VWF) which unfold in the shear forces of the microvasculature. HMW VWF multimers recruit platelets to the wounded endothelium. Unfolded HMW multimers consume circulating ADAMTS13, allowing for increased platelet binding to uncleaved HMW multimers downstream. In turn, activated platelets aggregate activating coagulation and forming microvascular thrombi. In high shear stress, schistocytes are formed as a result of RCB shearing while forced through small vessels with thrombi. High D-Dimers result from plasmin degradation of microthrombi.

**Table 3).** Clinical Laboratory Data within the First 72 Hours of Admission from Patients with COVID19 Stratified by ADAMTS13 Activity Level

| Characteristics |  | Low ADAMTS13 Activity<br>(<43%)<br>(n=33<sup>a</sup>) (median [IQR] or n (%)) | High ADAMTS13 Activity<br>(≥43%)<br>(n=69<sup>a</sup>) (median [IQR] or n (%)) | p |
| --- | --- | --- | --- | --- |
| Age |  | 71.0 [62.0, 80.0] | 68.0 [59.0, 79.0] | 0.31 |
| Sex (Male) |  | 22 (67) | 41 (59) | 0.50 |
| Race | Black or African-American | 7 (21) | 28 (40) | 0.09 |
|  | White | 1 (3) | 10 (15) | 0.10 |
|  | Other/Patient Declined | 25 (76) | 31 (45) | 0.003 |
| Ethnicity | Spanish/Hispanic/Latino | 16 (49) | 23 (33) | 0.21 |
| BMI (kg/m<sup>2</sup>) |  | 30.9 [26.2, 32.8] | 28.9 [26.9, 31.3] | 0.25 |
| Elixhauser Comorbidity Index |  | 4.0 [2.0, 6.0] | 4.0 [1.0, 6.0] | 0.71 |
| Mortality |  | 23 (70) | 28 (41) | 0.01 |
| Invasive Ventilator Use |  | 12 (36) | 11 (16) | 0.04 |
| In Vivo Thrombosis or Ex Vivo Clot |  | 5 (15) | 11 (16) | >0.99 |
| Vasopressor Use |  | 6 (18) | 10 (15) | 0.85 |
| Hemodialysis or CRRT Use |  | 5 (15) | 7 (10) | 0.52 |
| Anticoagulation Use<sup>b</sup> |  | 8 (24) | 9 (13) | 0.26 |
| Steroid Use<sup>c</sup> |  | 10 (30) | 19 (28) | 0.96 |
| Clinical Laboratory Values Measured at Time of ADAMTS13 Activity (units) [Reference Range] |  |  |  |  |
| ADAMTS13 Activity (%) [70-110] |  | 34.5 [26.8, 38.6] | 66.2 [50.9, 79.9] | <0.001 |
| Schistocyte Count (%) [<0.5] |  | 2.04 [1.05, 2.85] n=13 | 0.48 [0.18, 0.68] n=20 | 0.01 |
| Schistocyte >1% |  | 10 (30) | 1 (1) | <0.001 |
| Ghost Cell Count (%) |  | 0.39 [0.2, 0.55] n=13 | 0.27 [0.08, 2.77] n=20 | 0.07 |
| Lactate Dehydrogenase (U/L) [140-280] |  | 555.0 [417.0, 693.0] | 452.0 [283.0, 625.5] | 0.03 |
| Indirect Bilirubin (mg/dL) [0.2-0.8] |  | 0.25 [0.10, 0.40] | 0.25 [0.10, 0.40] | 0.61 |
| Hemoglobin (g/dL) [12-17.5] |  | 12.9 [11.2, 14.6] | 12.5 [10.4, 14.3] | 0.35 |
| Platelet Count (k/uL) [150-450] |  | 197.0 [149.0, 270.0] | 211.0 [145.0, 304.0] | 0.54 |
| Platelet Trajectory<sup>d</sup> | Decrease | 16 (49) | 9 (13) | <0.001 |
|  | Increase | 11 (33) | 37 (54) | 0.06 |
| Creatinine (mg/dL) [0.84-1.21] |  | 1.6 [0.8, 2.4] | 1.1 [0.9, 2.0] | 0.40 |
| Increased Creatinine Trajectory<sup>d</sup> |  | 14 (42) | 16 (23) | 0.08 |
| VWF Activity (Ristocetin) (%) [50-150] |  | 352.0 [225.0, 490.0] | 258.0 [200.0, 322.0] | 0.04 |
| VWF Antigen (%) [50-150] |  | 442.0 [282.0, 656.0] | 346.0 [256.0, 440.0] | 0.06 |
| D-Dimer (ug/mL) [<.27] |  | 2.64 [1.86, 13.98] | 1.96 [0.89, 6.78] | 0.09 |
| Fibrin Monomer (ug/mL) [<10] |  | 5.0 [5.0, 61.8] | 5.0 [5.0, 24.5] | 0.29 |
| Factor VIII Activity (%) [50-150] |  | 144.0 [112.0, 153.0] n=11 | 154.0 [98.0, 188.0] n=37 | 0.81 |
| HMW Multimer Fold Change from Normal<sup>e</sup> |  | 0.90 [0.68, 1.70] n=13 | 1.01 [0.87, 1.20] n=36 | 0.63 |
| Fibrinogen (mg/dL) [200-400] |  | 510.0 [377.5, 681.0] n=19 | 588.0 [458.8, 747.5] n=48 | 0.43 |
| Initial Clinical Laboratory Values Measured upon Admission (units) [Reference Range] |  |  |  |  |
| eGFR (mL/min/1.73m<sup>2</sup>) [90-120] |  | 39.0 [26.0, 83.0] | 56.0 [30.8, 83.4] | 0.73 |
| Aspartate Transaminase(IU/L) [8-48] |  | 53.5 [42.5, 106.8] | 44.0 [28., 69.0] | 0.06 |
| Mean Platelet Volume (fL) [7-12] |  | 11.2 [10.7, 11.6] | 10.7 [10.0, 11.8] | 0.15 |
| Lymphocyte (count) [1.5-4.5] |  | 0.8 [0.6, 0.9] | 1.1 [0.7, 1.5] | 0.02 |
| International Normalized Ratio [<1.1] |  | 1.1 [1.0, 1.3] | 1.2 [1.1, 1.3] | 0.38 |
| C-Reactive Protein (mg/L) [<10] |  | 17.0 [5.3, 27.1] | 13.7 [4.5, 20.4] | 0.19 |
| Troponin (ng/mL) [<0.04] |  | 0.04 [0.01, 0.08] | 0.01 [0.01, 0.04] | 0.05 |
| Pulse Oximeter (%) [95-100] |  | 92.0 [84.0, 96.0] | 96.0 [91.0, 99.0] | 0.03 |
| Diastolic Blood Pressure (mm Hg) [60-80] |  | 70.0 [46.0, 80.0] | 72.0 [61.0, 82.0] | 0.45 |

a. Unless otherwise stated
b. within 48 hours before clot or ADAMTS13 measurement
c. within 24 hours before ADAMTS13 measurement
d. Based on a change >10% over 1 week period
e. See figure 2
Abbreviations: IQR, interquartile range; BMI, body mass index; CRRT, Continuous Renal Replacement Therapies; ADAMTS13, a disintegrin and metalloproteinase with a thrombospondin type 1 motif, member 13; VWF, Von Willebrand Factor; HMW, High Molecular Weight; eGFR, Estimated Glomerular Filtration Rate

Severe thrombocytopenia at presentation was rare, with only one patient having a platelet count of 1 k/µl and an ADAMTS13 level of 118%. Although admission platelet count was not significantly different between patients with ADAMTS13 <43% vs. ≥43%, the trajectory (defined as a change >10% within 7 days) was significantly different. 16 (49%) patients admitted with ADAMTS13 <43% had a decreased in their platelets compared to 9 (13%) patients with ADAMTS13 ≥43%; p<0.001 (Table 3). The majority (16/25, 64%) of patients with a negative platelet trajectory died.

D-Dimer, FM, fibrinogen, and FVIII were not significantly different in patients with initial ADAMTS13 <43% compared to patients with ADAMTS13 ≥43%. Although a strong correlation was observed between initial D-Dimer and FM, no correlation was observed between initial D-Dimer and prothrombin time (PT), FVIII, platelet count, and hemoglobin (Supplemental Figure 6).

### ADAMTS13 levels <30% are not caused by immune mediated antibodies

To investigate the etiology of the decreased ADAMTS13, we assessed inhibitor status for the protease in all cases with an ADAMTS13 <30%, which is a routine cut off for further work up for antibody detection. 12% (22 of 181) of patients in our cohort had ADAMTS13 levels <30% with mild inhibition <40% (Table 4), but when these samples were tested for specific antibodies against ADAMTS13 by ELISA, none were found in any of these patients. Since IL-6 can inhibit ADAMTS13, we correlated ADAMTS13 with IL-6 levels;^16^ however, we did not observe a linear correlation between IL-6 and degree of ADAMTS13 inhibition (r=0.06). Likewise, ADAMTS13 level did not directly correlate with eGFR or AST. Assessment for dysfunctional ADAMTS13 was unrevealing: ADAMTS13 antigen levels were correspondingly low in the 9 patients in whom it was measured (0.1-0.4, normal range 0.6-1.6 UI/mL, data not shown).

**Table 4).**
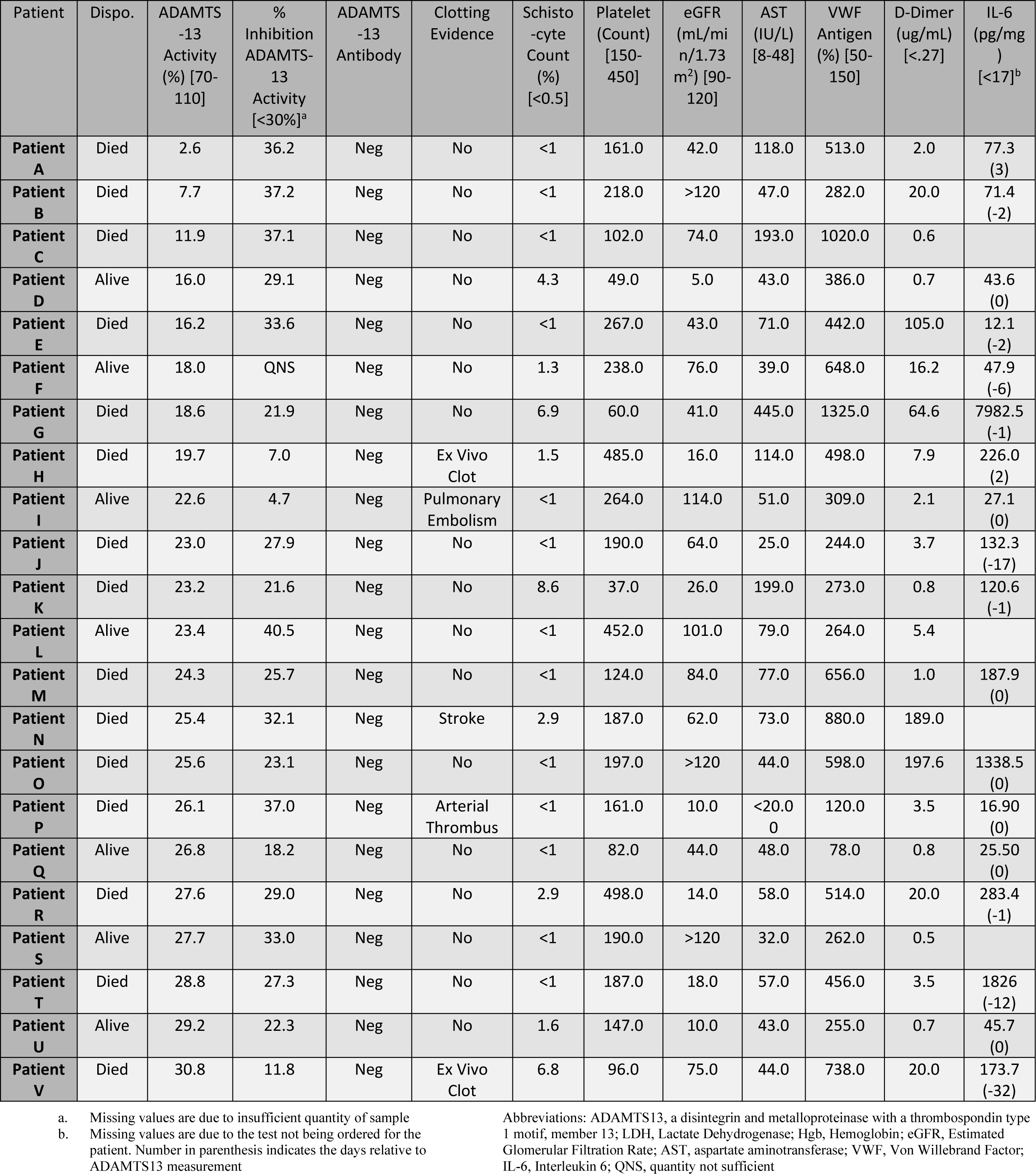
ADAMTS13 Activity Inhibitor, Kidney Function, Liver Function, and Immunological Analysis of Patients with very low (≤30) ADAMTS13 Activity

## Discussion

The main hypothesis of this retrospective study is that endothelial activation is associated with coagulation and microvascular thrombosis in COVID-19. We performed a balanced retrospective study of COVID-19 hospitalized patients with similar demographics and comorbidities and a wide range of D-Dimer levels to study how markers of endothelial activation correlate with coagulation, intravascular hemolysis and outcome. Indeed, we show a clear association of elevated VWF levels with high D-Dimer and FM levels. We also show that mild ADAMTS13 deficiency is common in COVID-19 inpatients.

Elevated VWF levels have been documented in COVID-19.^9,11,17^ Likewise, inflammatory markers such as CRP and IL-6 are known to be elevated in COVID-19. Thus, a potential explanation for elevated VWF levels in COVID-19 could be that this represents an acute phase response.^18,19^ However, the magnitude of increases in D-Dimer, FM, VWF cannot be explained solely by acute phase response and/or inflammation. In addition, ADAMTS13 is not expected to significantly decrease in acute inflammation, yet the majority of COVID-19 patients had decreased ADAMTS13, indicating a profound endothelial dysregulation or an intrinsic ADAMTS13 deficiency. Possible mechanisms of ADAMTS13 deficiency include decreased production, inhibition or consumption. 12% of patients in our cohort had ADAMTS13 activity levels less than 30% but none had detectable anti-ADAMTS13 antibodies. Many of these patients had increased IL-6 levels, but the IL-6 level did not correlate linearly with reduced ADAMTS13, thus favoring consumption or decreased production rather than inhibition. ADAMTS13 antigen levels were also reduced, but approximately 50% of patients with ADAMTS13 levels <30% had normal liver function, so production was not impaired in these patients. Consumption of ADAMTS13 due to excess of its substrate, VWF, or excess of plasmin, has been observed in sepsis, disseminated intravascular coagulopathies (DIC) and thrombotic microangiopathy (TMA) ^20-23^.

Thrombocytopenia is not common in COVID-19 and was not directly associated with low ADAMTS13 levels in our cohort.^24^ Also, lack of severe ADAMTS13 deficiency (only two patients had ADAMTS13 <10%) and lack of anti-ADAMTS13 antibodies in our patients excludes thrombotic thrombocytopenic purpura ^25^ and is more suggestive of secondary TMAs, which can be caused by viral infections.

TMA is defined by the triad of microangiopathic anemia, thrombocytopenia and end-organ damage.^26^ In our cohort we found evidence of microangiopathic anemia (schistocytes) and intravascular hemolysis (high LDH, indirect bilirubin) in the majority of the patients with low ADAMTS13. Among patients with low ADAMTS13, many had decreasing platelet trajectories and evidence of platelet consumption in peripheral blood smears (large immature platelets and platelet clumps). The correlation of schistocytes with markers of hemolysis (LDH, indirect bilirubin), and elevated VWF levels with a concomitant presence of decreased ADAMTS13 is highly suggestive of a thrombotic microangiopathy pattern. Also, the correlation of high D-Dimer and FM with LDH and the occasional finding of fibrin strands in peripheral blood smears suggests that high D-Dimer levels may be a direct product of small vessel thrombosis (arterial and venous), which have been documented in COVID-19 autopsies.^27-32^ Microvascular thrombosis leads to ischemic end-organ damage, most commonly affecting kidneys, but other organs can be affected in TMA. COVID-19 primarily manifests as respiratory failure, however, renal and cardiovascular complications are common in COVID-19. In our cohort approximately 40% of patients required ventilation, 20% of patients developed thrombosis, and 15% required hemodialysis, of which 28% developed *ex vivo* clots. We observed a trend of both *in vivo* and *ex vivo* thrombosis in cases with lower ADAMTS13 but did not reach a significance of p<0.05, probably due to the small sample size.

High D-Dimer, coagulation factor consumption, anemia and thrombocytopenia along with multiple organ damage can be seen in DIC.^33^ However, in our cohort high D-Dimer did not correlate with prolonged PT, and unlike DIC, COVID-19 patients presented with increasing fibrinogen, FVIII and FM along with D-Dimer (Supplemental Figure 6). Furthermore, high D-Dimer did not correlate with decreasing hemoglobin or platelets, altogether suggesting a TMA-like pattern as opposed to DIC.

Approximately 60% of patients that developed thrombosis were on anticoagulation at least 48 hours before clot detection. However, patients receiving anticoagulation had higher LDH and VWF levels, suggesting these patients were sicker (Supplemental Figure 5). Many other studies have shown that despite anticoagulation, certain COVID-19 patients still thrombose.^34^ Anticoagulation alone is not an effective treatment for TMA.^35^ Anecdotal evidence suggest that TMA treatments may be effective in COVID-19. Eculizumab binds C5, inhibiting the terminal complement complex, and has been shown to be effective in treating COVID-19 in several case reports. ^36-38^ Although we did not measure complement levels in our cohort, as serum samples were not preserved, we noticed ghost cells in several cases, which suggest complement activation on red blood cells.^39^ A nanobody, caplacizumab, that inhibits the binding of VWF to gp-1b on platelets has been shown effective to treat TMAs. ^40^ ADAMTS13 replacement via plasma exchange is a standard TTP treatment. ^41^ Although, the main rationale of convalescent plasma (CP) treatment is to provide passive immunity to acutely ill COVID19 patients, replacement of ADAMTS13 and other plasma proteins can possibly contribute to benefits attributed to CP. ^42-44^

An advantage of our study is that cases were selected based on a repository of frozen plasma, and thus multiple tests with serial dilutions were performed, allowing us to accurately correlate D-Dimer, FM, VWF activity, VWF antigen, VWF multimers, FVIII, and ADAMTS13 levels, all derived from the same samples. In addition, serial dilutions of samples that reach the upper limit of detection allow us to accurately measure the actual levels of high D-Dimer, FMs and VWF levels. Herein, we showed cases with unprecedented levels of VWF >1000%, FM > 2000 μg/ml and D-Dimer > 300 μg/ml FEU.

Like any other retrospective studies, limitations include intrinsic confounders and bias. Choosing samples from a limited repository bank could create bias. We tried to compensate by randomly selecting a balanced cohort with equal distribution of survivors and non-survivors and similar demographics. We could only demonstrate correlations but no causality. Major confounders include: a wide spectrum of disease severity at presentation, and over imposed sepsis. Thus, we cannot exclude the possibility that low ADAMTS13 is simple a passive biomarker and an indirect consequence of disease severity. Therefore, prospective randomized clinical studies are needed to determine the relationship and causality between ADAMTS13 levels, complement, endothelial, and coagulation activation and to study the efficacy of TMA treatments in treating COVID-19.

In summary, we present the most comprehensive and largest study to date analyzing correlations of D-Dimer levels with VWF activity/antigen, size of VWF multimers, ADAMTS13 levels, markers of intravascular hemolysis and smear pathology in hospitalized COVID-19 patients. Low ADAMTS13 and increased schistocytes on admission correlated with mortality. Thus, in addition to D-Dimer, presence of schistocytes on admission may warrant a work up for TMA, including ADAMTS13 levels, since this group may require different/additional therapy.

## Data Availability

Data is available upon email request

## Acknowledgements

We will like to acknowledge the MSTP Training grant for partial support of Joseph Sweeney’s effort, T32GM007288. We will like to thank Drs. Henny Billet, Eran Billen and Monika Paroder for their time and suggestions to improve the presentation and style of this study. We will like to thank Albert Einstein College of Medicine medical student Maxwell Roth for his digital art contributions. We will like to acknowledge the coagulation lab manager, Julissa Pena and lab technologists Mayra Almonte, Leli Soriano and Geovanna Cruz for their dedication handling and processing clinical specimens during the COVID crisis. Finally, we will like to express our support to all COVID-19 patients and their families.

Morayma Reyes Gil and Joseph Sweeney had full access to all the data in the study and take responsibility for the integrity of the data and the accuracy of the data analysis and include this in the Acknowledgment section of the manuscript.

## Authorship and Competing interest

MRG designed research, performed research, analyzed and interpreted data, wrote manuscript.

JMS performed research, analyzed and interpreted data, and helped in manuscript writing.

MB analyzed and interpreted data, and helped in manuscript writing.

GJK performed research and analyzed data.

JDG performed chart review.

SR performed chart review.

All authors declare no conflict of interest.

## Supplementary Figures and Tables

**Supplementary Figure 1).**
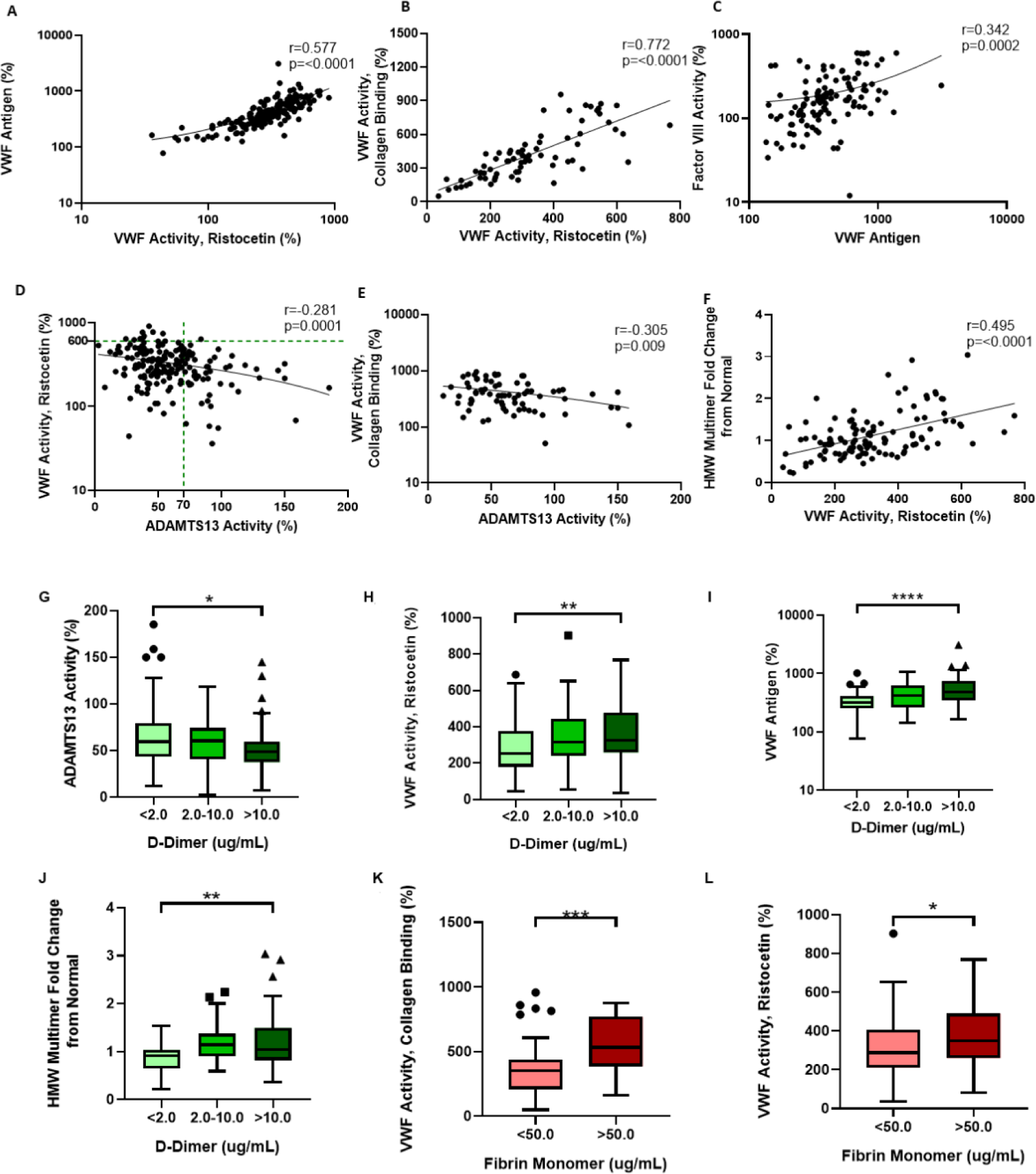
Correlations amongst various endothelial activation and coagulation parameters. The Pearson’s coefficient (r), p-value, and trendline is shown for graphs A-F. All 181 patients are represented unless otherwise stated. A) Scatter plot showing positive correlation between VWF Antigen and VWF Ristocetin activity. B) Scatter plot showing positive correlation between VWF Collagen Binding activity and VWF Ristocetin activity. This only includes patients for whom an ELISA for VWF Collagen Binding activity was completed (n=72) C) Scatter plot showing slight positive correlation between Factor VIII activity and VWF Antigen (n=116) D) Scatter plot showing negative correlation between VWF Ristocetin activity and ADAMTS13 Activity. Almost all (10/11) VWF activity levels greater than 600% occur in patients with ADAMTS13 levels less than 70%. E) Scatter plot showing negative correlation between VWF Collagen Binding activity and ADAMTS13 Activity (n=72). F). Scatter plot showing positive correlation between VWF Ristocetin activity and the fold change of each patient’s HMW multimer size compared to the HMW multimer size of the normal pooled plasma control. This only includes patients for whom multimer western blots were ran (n=115). G-L. Within each box plot, the horizontal line indicates the median, the outside bars indicate the 25^th^ and 75^th^ percentile, individual dots indicate outlier points, and asterisk represent the p-value from a one-way ANOVA (if three values), or two tailed t-test (if two values). The asterisk indicates significance as follows: *p < 0.05, **p < 0.01, ***p < 0.001, and **** P<0.0001. The Box Plot shows G) ADAMTS13 Activity (n=181) H) VWF Ristocetin activity (n=181) I) VWF Antigen (n=181) J) fold change in High Molecular Weight Multimer compared to normal (n=115), stratified by low (<2ug/mL), medium (2.0-10.0 ug/mL), or high (>10ug/mL) D-dimer concentration. Generally, each lab parameter became more abnormal in the medium and high D-Dimer stratification compared to the low stratification. The Remaining box plots show K) VWF Collagen Binding Activity (n=72) and L) VWF Ristocetin activity (n=181), stratified by low (<50ug/mL) or high (>50ug/mL) Fibrin Monomer concentration. Generally, each lab parameter was more abnormal in the high Fibrin Monomer stratification compared to the low stratification.

**Supplementary Figure 2).**
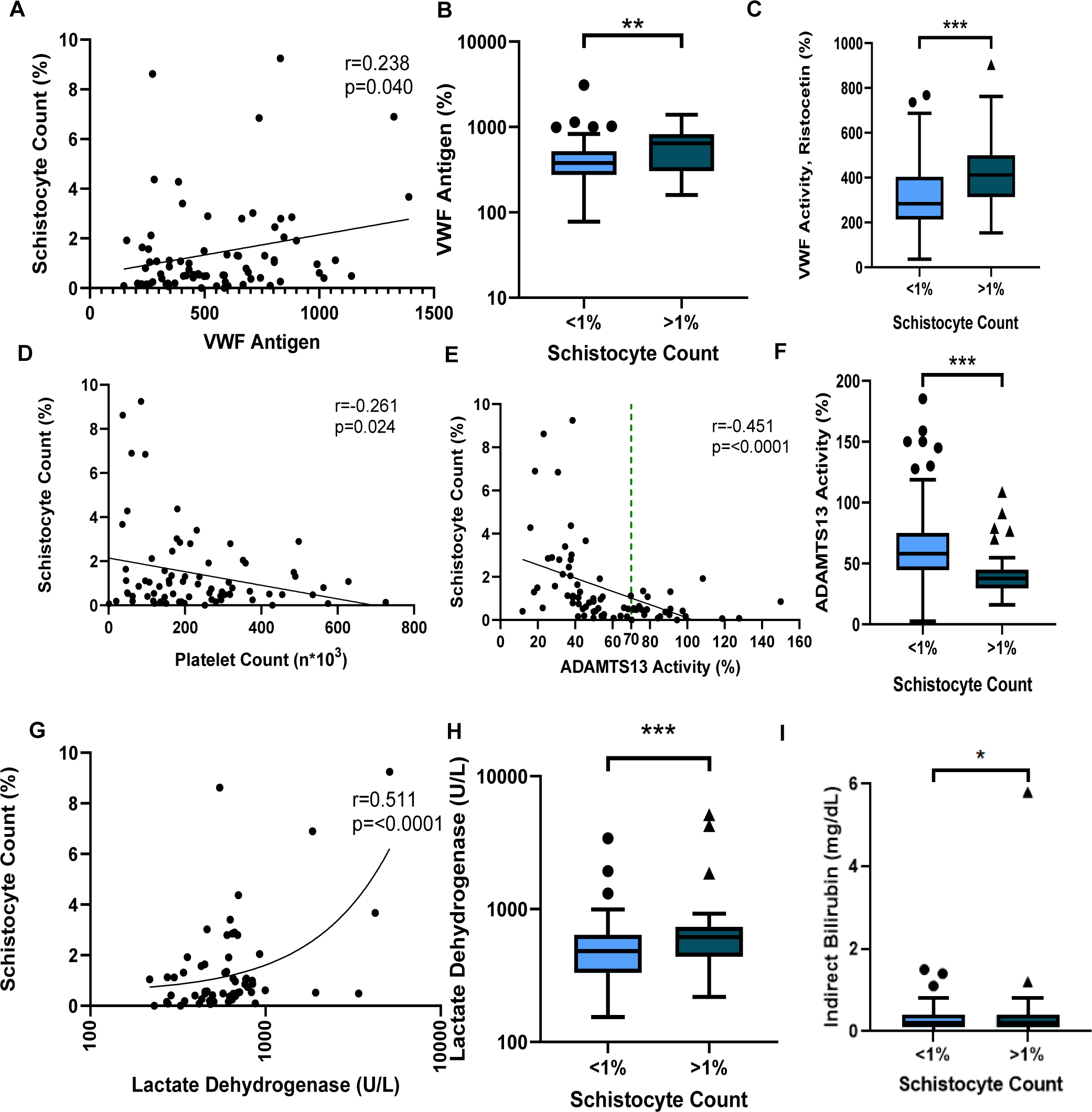
Correlation of various hemolysis, endothelial activation, and coagulation parameters to Schistocyte Count. For each scattter plot (A, D, E G), the Pearson’s coefficient (r), p-value, and trendline is shown. Only patients for whom a CBC was flagged as abnormal within three days of when the sample was taken and therefore could be specifically quantified are included in the scatter plots (n=73). A) Scatter plot showing positive correlation between the Schistocyte count and VWF antigen. D) Scatter plot showing negative correlation between the Schistocyte count and Platelet count. All cases of schistocyte counts greater than 4% occurred in patients with a platelet count less than 200,000/ml E) Scatter plot showing negative correlation between the Schistocyte count and ADAMTS13 activity. All cases of schistocyte counts greater than 2% occurred in patients who had ADADMTS13 activity less than normal 70% (dotted green line). G) Scatter plot showing positive correlation between the Schistocyte count and lactate dehydrogenase. For each box plot (B, C, F, H, I), the horizontal line indicates the median, the outside bars indicate the 25^th^ and 75^th^ percentile, individual dots indicate outlier points, and the asterisk represent the p-value from a two tailed t-test. The asterisk indicates significance as follows: *p < 0.05, **p < 0.01, ***p < 0.001, and **** P<0.0001. All patients are included in the box plots (n=181) unless otherwise noted, with patients without an abnormal CBC flag within three days of the sample assumed to have <1% schistocyte count (see methods). The box plots show B) VWF antigen C) VWF Ristocetin activity F) ADAMTS13 H) Lactate Dehydrogenase (n=158) and I) Indirect Bilirubin levels (n=171) stratified by low (<1%) or high (>1%) schistocyte count.

**Supplementary Figure 3).**
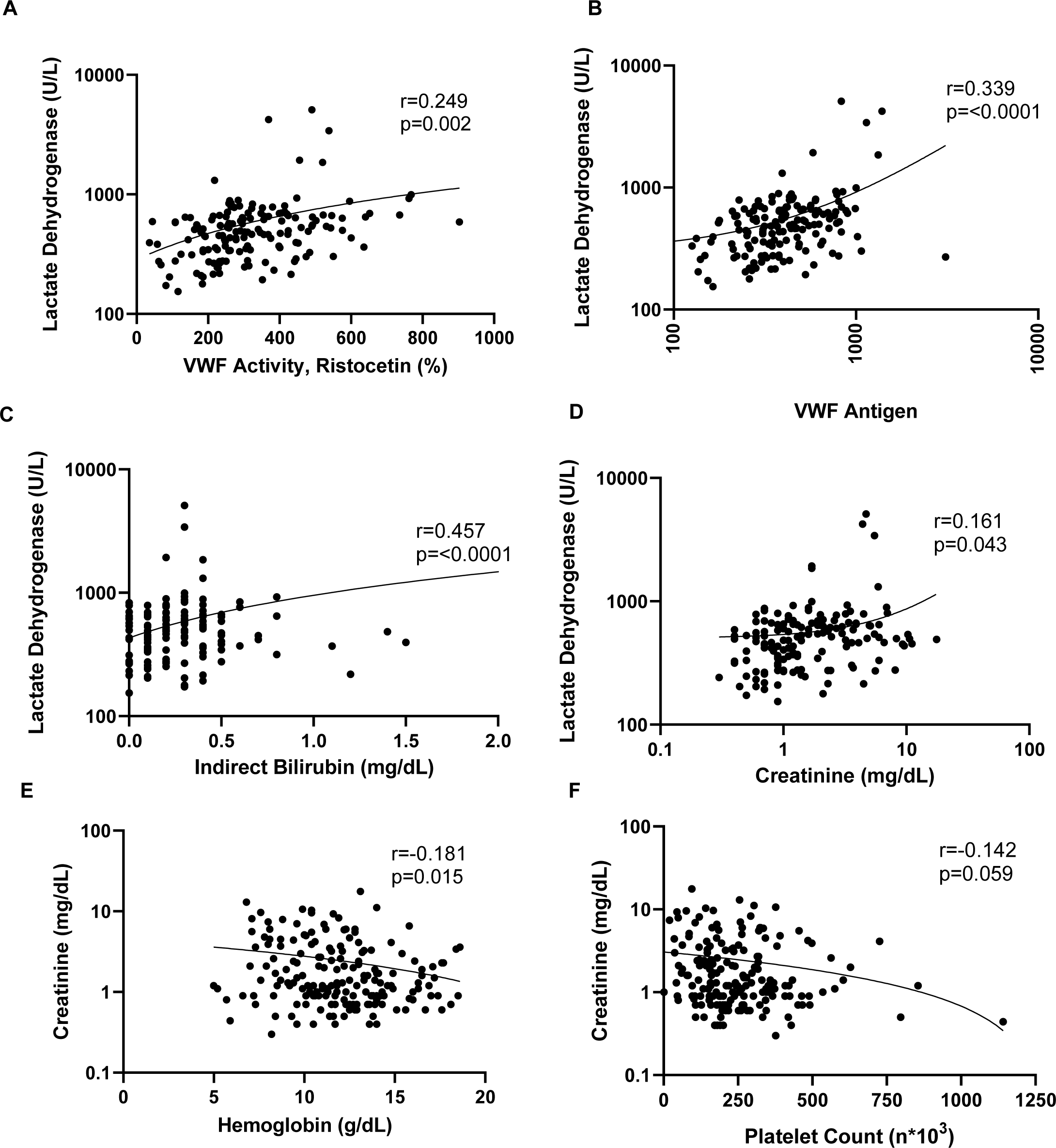
Correlations amongst various lab values measured to assess hemolysis and/or coagulopathies. The Pearson’s coefficient (r), p-value, and trendline is shown for each graph. All 181 patients are represented in each plot unless otherwise stated. A) Scatter plot showing positive correlation between lactate dehydrogenase level and VWF Ristocetin activity (n=158) B) Scatter plot showing positive correlation between lactate dehydrogenase level and VWF antigen (n=158). C) Scatter plot showing positive correlation between lactate dehydrogenase level and indirect bilirubin. Only cases for which an LDH and bilirubin measurements were taken within 48 hours of the sample are included (n=158) D) Scatter plot showing positive correlation between lactate dehydrogenase level and creatinine level (n=158) E) Scatter plot showing negative correlation between creatinine level and hemoglobin level F) Scatter plot showing negative correlation between creatinine level and platelet count.

**Supplementary Table 1).** Thrombotic Events and Anticoagulation Treatment of 181 Patients with COVID19 Stratified by ADAMTS13 Activity Level

| Characteristics, median [IQR] or n (%) | Low ADAMTS13 Activity<br>(<70%) (n=129<sup>a</sup>) | Normal ADAMTS13<br>Activity (>70%) (n=52<sup>a</sup>) | p |
| --- | --- | --- | --- |
| ADAMTS13 Activity (%) [70-110] | 46.1 [36.7, 55.4] | 88.71 [76.3, 100.2] | <0.001 |
| Schistocyte Count (%) [<0.5] | 1.06 [0.52, 2.54] | 0.48 [0.16, 0.59] | <0.001 |
| Age | 68.0 [59.0, 78.0] | 64.0 [49.8, 73.0] | 0.02 |
| Sex (M) | 77 (59.7) | 29 (56) | 0.63 |
| Mortality | 71 (55.0) | 19 (37) | 0.02 |
| Continuous Renal Replacement Therapies | 19 (14.7) | 6 (12) | 0.75 |
| Hemodialysis Use | 22 (17.1) | 7 (14) | 0.71 |
| **Thrombotic or Clotting Event within entire Hospital Admission, n (%)** |  |  |  |
| Thrombosis | 23 (17.8) | 13 (25) | 0.37 |
| • Deep Venous Thrombosis | 14 (10.9) | 10 (19) | 0.21 |
| • Pulmonary Embolism | 3 (2.3) | 1 (2) | 1.000 |
| • Arterial Thrombosis | 3 (2.3) | 2 (4) | 0.63 |
| • Stroke | 2 (1.6) | 0 (0) | 1.000 |
| Ex Vivo Clotting | 10 (7.8) | 1 (2) | 0.18 |
| **Thrombotic or Clotting Event within 7 Days of ADAMTS13 Activity Measurement, n (%)** |  |  |  |
| Thrombosis | 16 (12.4) | 4 (8) | 0.44 |
| • Deep Venous Thrombosis | 10 (7.8) | 3 (6) | 0.76 |
| • Pulmonary Embolism | 1 (0.8) | 0 (0) | 1.000 |
| • Arterial Thrombosis | 3 (2.3) | 1 (2) | 1.000 |
| • Stroke | 2 (1.6) | 0 (0) | 1.000 |
| Ex Vivo Clotting | 7 (5.4) | 0 (0) | 0.20 |
| **Anticoagulation<sup>b</sup>, n (%)** |  |  |  |
| None | 80 (62.1) | 34 (65) | 0.67 |
| Prophylactic | 26 (20.2) | 12 (23) | 0.81 |
| • Heparin | 9 (7.0) | 1 (2) | 0.29 |
| • Enoxaparin | 10 (7.8) | 4 (8) | 1.000 |
| • Apixaban | 7 (5.4) | 7 (14) | 0.13 |
| Therapeutic | 23 (17.8) | 6 (12) | 0.41 |
| • Heparin | 1 (0.8) | 1 (2) | 0.49 |
| • Enoxaparin | 4 (3.1) | 0 (0) | 0.58 |
| • Apixaban | 9 (7.0) | 3 (6) | 1.000 |
| • Bivalirudin | 8 (6.2) | 2 (4) | 0.73 |
| • Warfarin | 1 (0.8) | 0 (0) | 1.000 |
| **Anticoagulation before Thrombosis or Ex Vivo Clot<sup>c</sup>, n (%)** |  |  |  |
| None | 13/30 (43) | 5/13 (38.5) | 1.000 |
| Prophylactic | 4/30 (13) | 4/13 (31) | 0.22 |
| Therapeutic | 11/30 (37) | 4/13 (31) | 1.000 |
| Change of Anticoagulation after | 19/30 (63) | 11/13 (85) | 0.30 |

a. Unless otherwise stated
b. all patients within 48 hours before clot or ADAMTS13 measurement
c. Out of patients who experienced thrombosis or an ex vivo clot; 48 hours before clot
Abbreviations: IQR, interquartile range; ADAMTS13, a disintegrin and metalloproteinase with a thrombospondin type 1 motif, member 13

**Supplementary Figure 4).**
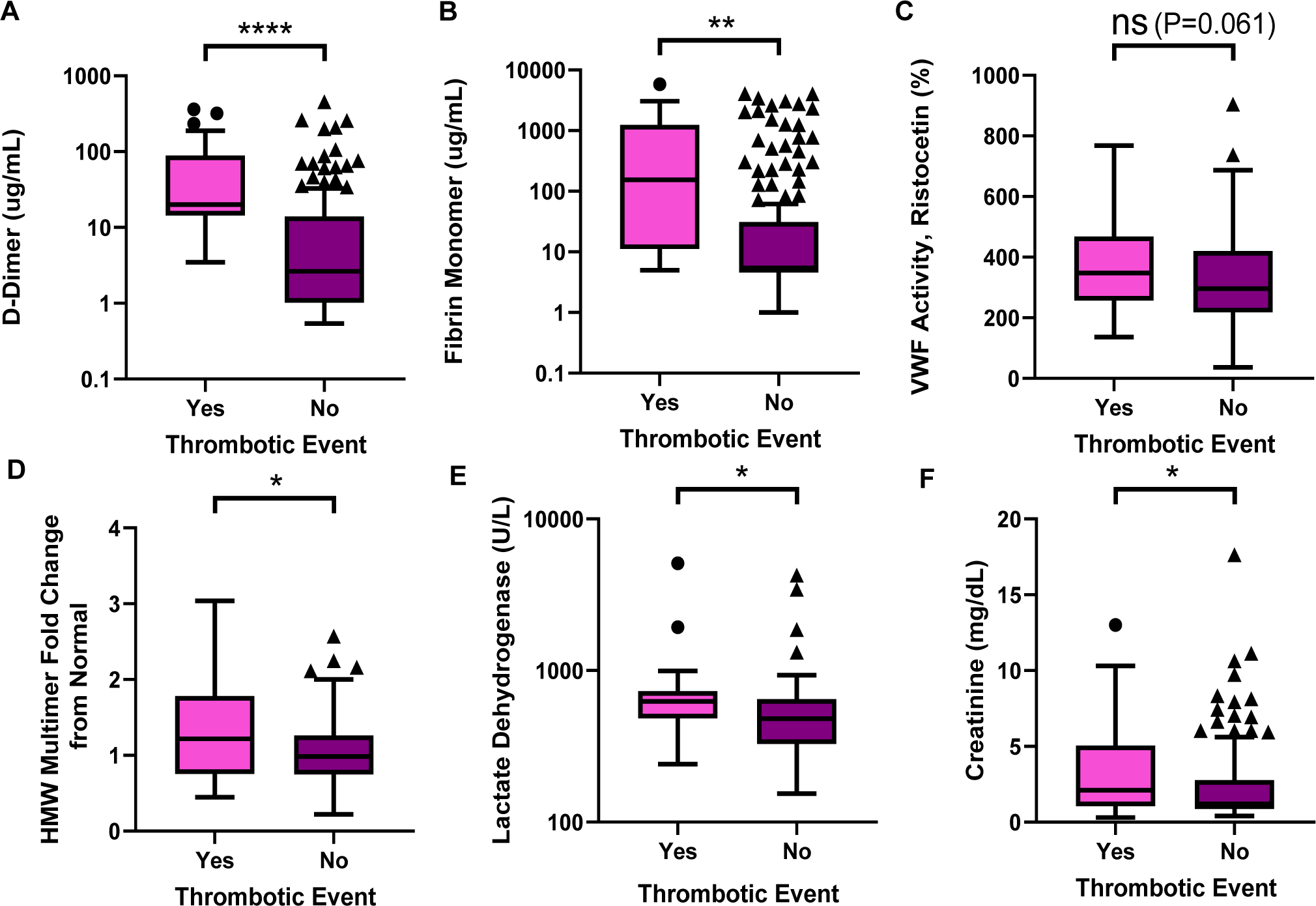
Markers of coagulation, endothelial activation, or hemolysis stratified by the occurrence of a thrombotic event. We considered a thrombotic event to be either an occurrence of in vivo thrombosis if it was documented with radiographic imagining, or an ex vivo clot if it was reported in the patient’s chart. All events within 7 days of the blood sample we used to measure the markers of coagulation and hemolysis were considered. Within each box plot, the horizontal line indicates the median, the outside bars indicate the 25^th^ and 75^th^ percentile, individual dots indicate outlier points, and asterisk represent the p-value from a two tailed t-test. The asterisk indicates significance as follows: *p < 0.05, **p < 0.01, ***p < 0.001, and **** P<0.0001. The Box Plots show A) D-Dimer B) Fibrin monomer C) VWF Ristocetin activity D) fold change of HMW multimer size compared to that of normal pooled plasma E) Lactate dehydrogenase or F) Creatinine level stratified by a thrombosis or clotting event within 7 days of the sample.

**Supplementary Figure 5).**
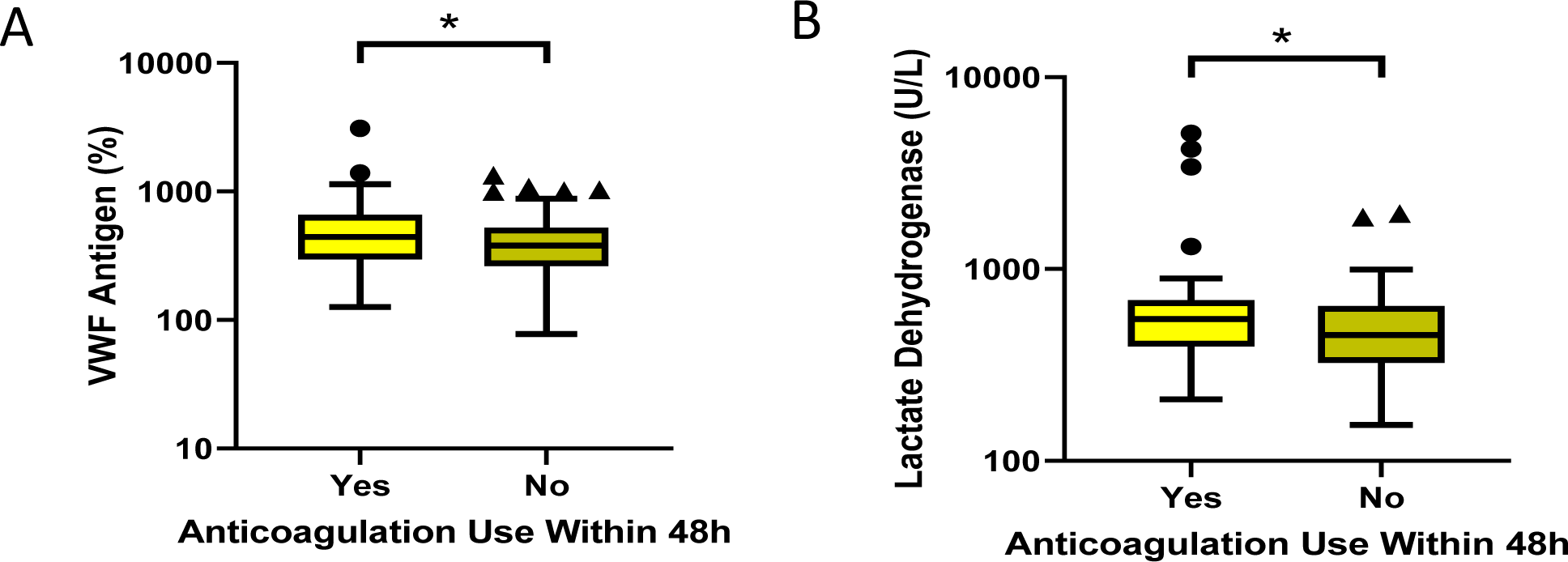
VWF antigen and Lactate dehydrogenase stratified by anticoagulation use. Within each box plot, the horizontal line indicates the median, the outside bars indicate the 25^th^ and 75^th^ percentile, individual dots indicate outlier points, and asterisk represent the p-value from a two tailed t-test. The asterisk indicates significance as follows: *p < 0.05, **p < 0.01, ***p < 0.001, and **** P<0.0001. The Box Plots show A) VWF antigen and B) Lactate dehydrogenase stratified by anticoagulation use. A patient was considered to be on anticoagulation medication if it was administered at least 48 hours prior to when the sample was taken.

**Supplementary Figure 6).**
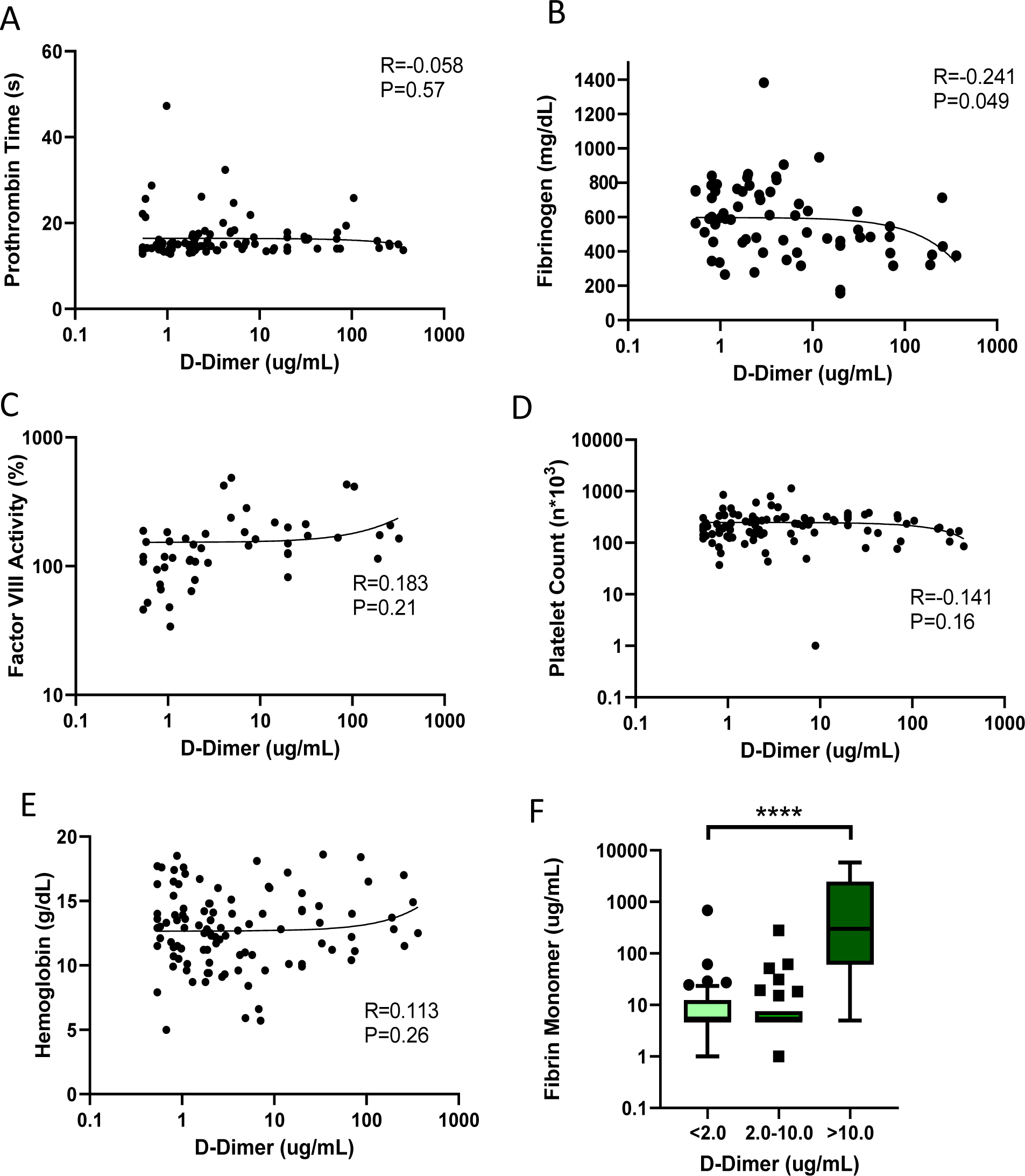
Correlation of D-Dimer with other classic markers of Disseminated intravascular coagulation (DIC) within 72 hours of admission. The Pearson’s coefficient (r), p-value, and trendline is shown for each graph. All 102 patients for whom an ADAMTS13 measurement was taken within 72 hours of admission are represented unless otherwise stated. A) Scatter plot showing no significant correlation between prothrombin time and D-Dimer B) Scatter plot showing slight negative correlation between fibrinogen and D-Dimer (n=67) C) Scatter plot showing no significant correlation between factor VIII activity and D-Dimer (n=48) D) Scatter plot showing no significant correlation between platelet count and D-Dimer E) Scatter plot showing no significant correlation between hemoglobin and D-Dimer F) Box plot of fibrin monomer stratified by low (<2ug/mL), medium (2.0-10.0 ug/mL), or high (>10ug/mL) D-dimer concentration. The horizontal line indicates the median, the outside bars indicate the 25^th^ and 75^th^ percentile, individual dots indicate outlier points, and asterisk represent the p-value from a one-way ANOVA. Four asterisks indicates the p value is <0.0001.

